# Seroprevalence of the hepatitis E virus among blood donors in the Qassim Region, Saudi Arabia

**DOI:** 10.1101/2019.12.19.19015412

**Authors:** Bader Y. Alhatlani, Waleed A. Aljabr, Mohammed S. Almarzouqi, Sami M. Alhatlani, Rayan N. Alzunaydi, Abeer S. Alsaykhan, Sulaiman H. Almaiman, Ahmed A. Aleid, Ammar H. Alsughayir, Yara E. Bishawri, Abdulrahman A. Almusallam

## Abstract

The aim of this study was to evaluate the seroprevalence of hepatitis E virus (HEV), a major public health issue worldwide with the potential for transmission via blood transfusion, in blood donors in the Qassim Region, Saudi Arabia. Serum samples (n = 1,078) were collected from volunteer blood donors from January to April 2019 and tested for the presence of anti-HEV IgG and IgM by indirect enzyme-linked immunosorbent assays. Overall, the seroprevalence of anti-HEV IgG and IgM among blood donors was 5.7% and 1.3%, respectively. Additionally, the seropositive rates of anti-HEV IgG and IgM were significantly higher in non-Saudi donors (22.1% and 7.8%) than in Saudi donors (3% and 0.2%). The seroprevalence of anti-HEV IgG increased with age; however, there was no correlation between gender and anti-HEV IgG and/or IgM. The seroprevalence of HEV among blood donors in the Qassim Region was lower than previous estimates for other regions of the country. Further studies covering a wider geographical area are needed to validate and expand the findings and to determine the importance of HEV screening in the region.

## Introduction

Hepatitis E virus (HEV), belonging to the *Orthohepevirus* genus in the family *Hepeviridae*, causes liver diseases in humans [1]. The virus was first described in the early 1980s as a non-A and non-B hepatitis virus and was subsequently cloned in 1991 [2,3]. HEV contains a small nonenveloped, positive-sense single-stranded RNA genome approximately 7,200 nucleotides long [4]. HEV is an etiological agent of hepatitis E in humans. Similar to hepatitis A, the vast majority of HEV infections are asymptomatic (especially in children) or cause self-limiting acute liver inflammation, which can resolve within a few weeks without the need for specific treatment [4]. However, immunosuppressed individuals, organ-transplant recipients, hemodialysis patients, and pregnant women are at a high risk of developing life-threatening diseases, including chronic hepatitis and acute liver failure, after infection with HEV [5,6]. It has been estimated that the mortality rates for HEV infections in pregnant women and young people are about 20% and 3%, respectively [7]. The incubation period, which occurs during the prodromal phase, can vary from 2 to 8 weeks, and common symptoms of HEV infection during this period are usually nonspecific and include fever, nausea, vomiting, and malaise [8]. HEV is now recognized as a major public health issue, causing over 20 million infections every year worldwide and accounting for approximately 70,000 deaths [1]. Currently, there are eight known HEV genotypes from only one serotype that can infect humans and other animal taxa (HEV-1 to HEV-8), of which HEV-1 and HEV-2 are restricted to humans and are associated with most outbreaks in developing countries in parts of Asia, Central America, and Africa [9]. Genotypes 3 and 4 are typically identified in developed countries, including the USA, New Zealand, Japan, and some countries in Europe, and can be isolated from a broader range of taxa, including humans, pigs, deer, and rabbits. Genotypes 5 and 6 are found in wild boars [8,10]. More recently, a new genotype of camelid HEV (HEV-7) was isolated from dromedary camels in the UAE and some African countries, including Sudan and Egypt [11,12].

Fecal-oral transmission is considered the main route of HEV transmission, but other transmission routes have been suggested. This includes person-to-person transmission, such as vertical transmission (mother-to-infant) during delivery. In particular, blood transfusion transmission has become one of the main routes, especially in some low-income countries in Asia [4]. Several studies have reported transfusion-transmitted HEV from blood components in many industrialized countries, including European countries, Australia, and the United States [13-18]. In the Gulf and other neighboring countries, few studies have evaluated the HEV seroprevalence in healthy blood donors. A recent study has indicated that the HEV seroprevalence in Qatar is high among blood donors, i.e., approximately 21% [19]. To the best of our knowledge, the first study of HEV in Saudi Arabia was conducted in 1994 in Riyadh and Gizan, with anti-HEV antibody detection frequencies of 8.4% and 14.9%, respectively [20]. Two additional studies of HEV in Saudi Arabia reported seroprevalences in blood donation samples of 18.7% and 16.9% in Makkah and Jeddah, respectively [21,22]. In most countries, screening is not available for HEV in blood donors. However, in the Netherlands, screening was introduced in 2017, and the United States is now considering HEV screening for blood donors [4]. In this study, we estimated the HEV seroprevalence among blood donors in the Qassim Region, Saudi Arabia. These results provide a basis for evaluating whether routine screening is necessary in Saudi Arabia.

## Materials and methods

### Ethics statement and sample collection

From January to April 2019, 1,078 whole blood samples were collected from volunteer blood donors at the Blood Donor Unit at King Saud Hospital, Unayzah, Saudi Arabia. All study participants provided written informed consent, and the study design was approved by the local research Ethical Committee of the General Directorate of Health Affairs at Qassim Province (approval number: 1441-225162).

### Serological testing

Serum samples were tested for the presence of anti-HEV IgG and IgM antibodies using commercial HEV Enzyme-linked Immunosorbent Assay (ELISA) Kits (Fortress Diagnostics, Antrim, UK) according to the manufacturer’s instructions. The sensitivity and specificity of the assays are 99.5% and 99.6%, respectively [23]. Samples were tested in duplicate and samples yielding borderline results were retested in duplicate to confirm the initial results. Only IgG-positive samples were tested for the presence of anti-HEV IgM.

### Statistical analyses

The Chi-square test and Fisher’s exact text with the Freeman-Halton extension were used to evaluate the associations between the demographic characteristics of the participants and HEV seroprevalence. All statistical analyses were performed using an online statistical calculator at: (https://www.socscistatistics.com); a two-tailed *p*-value of 0.05 was considered significant.

## Results

Whole blood samples (n = 1,078) were collected from blood donors at King Saud Hospital in the Qassim Region, Saudi Arabia. General characteristics of the blood donors are summarized in Table 1. The study population included 1,002 (93%) men and 76 (7%) women, with 924 (85.7%) Saudis and 154 (14.3%) non-Saudis with different nationalities. The age of participants ranged from 18 to 73 years (mean ± SD, 34.5 ± 10.3 years); 461 donors (42.8%) were aged 25 to 34 years, 202 donors (18.7%) were younger than 25 years, and 61 donors (5.7%) were older than 55 years (Table 1).

**Table 1.** Characteristics of the study population and anti-HEV IgG results.

| <u>Gender</u> | <u>Total (n = 1,078)</u> | <u>HEV IgG-positive</u> | <u>HEV IgG-negative (n</u> | <u>p-value</u> |
| --- | --- | --- | --- | --- |
|  | 76 (7%) | (n = 61) | = 1,017) |  |
| Female | 1,002 (93%) | 2 (2.6%) | 74 (97.4%) | 0.24 |
| Male |  | 59 (5.9%) | 943 (94.1%) |  |
| <u>Nationality</u> |  |  |  |  |
| Saudi | 924 (85.7%) | 27 (3%) | 897 (97%) | <0.001 |
| Non-Saudi | 154 (14.3%) | 34 (22.1%) | 120 (77.9%) |  |
| <u>Age group (years)</u> |  |  |  |  |
| <25 | 202 (18.7%) | 2 (1%) | 200 (99%) | 0.01 |
| 25–34 | 461 (42.8%) | 28 (6%) | 433 (94%) |  |
| 35–44 | 247 (22.9%) | 16 (6.5%) | 231 (93.5%) |  |
| 45–54 | 107 (9.9%) | 11 (10.3%) | 96 (89.7%) |  |
| ≥55 | 61 (5.7%) | 4 (6.5%) | 57 (93.5%) | |

In total, 61 of the 1,078 blood samples (5.7%) were positive for anti-HEV IgG, including 2 samples from women (2.6%) and 59 from men (5.9%), with no significant difference between males and females (*p* = 0.24). However, the anti-HEV IgG seroprevalence was significantly higher in non-Saudis (22.1%) than in Saudis (3%) (Table 1, *p* < 0.001). We also analyzed the same blood donor samples for the seroprevalence of anti-HEV IgM. Our results indicated that 14 of the 1,078 serum samples (1.3%) were positive for anti-HEV IgM (Table 2). Again, the seroprevalence of anti-HEV IgM was significantly higher in blood samples from non-Saudis (7.8%) than in those from Saudi donors (0.2%) (Table 2, *p* < 0.001). No significant differences were found in the frequencies of anti-HEV IgM-positive samples between women and men (*p* = 0.99).

**Table 2.** Characteristics of the study population and anti-HEV IgM results.

| <u>Gender</u> | <u>Total</u><br><u>(n = 1,078)</u> | <u>HEV IgM-positive</u><br><u>(n = 14)</u> | <u>HEV IgM-negative (n =</u><br><u>1,064)</u> | <u>p-value</u> |
| --- | --- | --- | --- | --- |
| Female | 76 (7%) | 1 (1.3%) | 75 (99%) | 0.99 |
| Male | 1,002 (93%) | 13 (1.3%) | 989 (99%) |  |
| <u>Nationality</u> |  |  |  |  |
| Saudi | 924 (85.7%) | 2 (0.2%) | 922 (99.8%) | <0.001 |
| Non-Saudi | 154 (14.3%) | 12 (7.8%) | 142 (92.2%) |  |
| <u>Age group (years)</u> |  |  |  |  |
| <25 | 202 (18.7%) | 2 (1%) | 200 (99%) | 0.72 |
| 25–34 | 461 (42.8%) | 6 (1.3%) | 455 (98.7%) |  |
| 35–44 | 247 (22.9%) | 5 (2%) | 242 (98%) |  |
| 45–54 | 107 (9.9%) | 1 (1%) | 106 (99%) |  |
| ≥55 | 61 (5.7%) | 0 (0%) | 61 (0%) |  |

Furthermore, we found a significantly higher rate of HEV IgG-positive samples for donors between 45 to 54 years old (10.3%, *p* = 0.01) than for other age groups, while donors younger than 25 years old had only an anti-HEV IgG seropositive rate of 1% (Fig 1). The other age groups had similar anti-HEV IgG seropositive rates, and there was no significant association between age groups and anti-HEV IgM seropositivity (*p* = 0.72).

**Figure 1.**
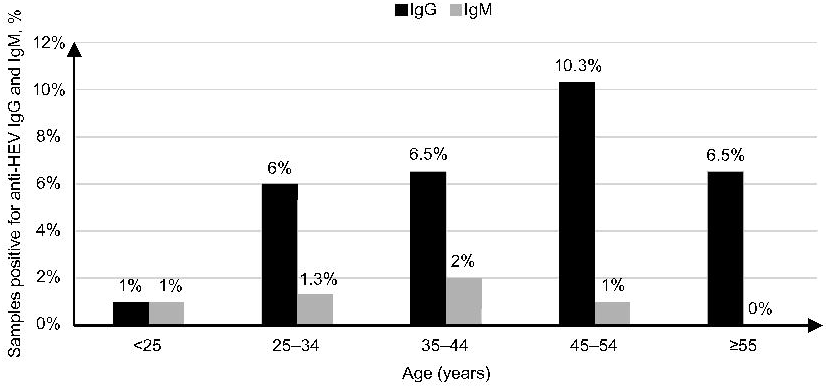
Age distributions of 1,078 blood donors positive for anti-HEV IgG and/or IgM. Of the 14 anti-HEV IgM-positive serum samples, only two samples tested positive for anti-HCV antibodies, both of which were from male non-Saudi donors. There was no significant association between anti-HEV and anti-HCV. In addition, none of the samples were positive for anti-HBsAg, anti-HIV, and anti-HTLV I/II.

## Discussion

HEV is a transfusion-transmissible virus, as evidenced by its detection in blood donors in both developed and developing countries [24,25]. In the current study, we screened 1,078 serum samples from blood donors in the Qassim Region, Saudi Arabia for anti-HEV IgG and IgM antibodies. This is the largest study of the seroprevalence of HEV in Saudi Arabia to date with respect to the number of samples obtained from blood donors. Our results indicated that the seroprevalence of HEV in blood donors is 5.7% for IgG and 1.3% for IgM. In addition, we detected a significant difference between Saudis (3%) and non-Saudis (22.1%) in the seroprevalence of anti-HEV IgG. The seroprevalence of anti-HEV IgM was also significantly higher among non-Saudi blood donors (7.8%) than in Saudi blood donors (only 0.2%). Moreover, we found an increase in the rate of anti-HEV IgG with age. The highest rate of positive anti-HEV IgG results was observed in the 45- to 54-year-old group and the lowest rate was obtained for donors younger than 25 years (Fig 1). These results are consistent with those of a previous study suggesting that age is a risk factor for HEV [26]. The anti-HEV IgM seroprevalence was comparable in all age groups other than donors aged over 55 years who tested negative for IgM. Some studies have suggested that the HEV seroprevalence rate increases with age, while other studies have reported that the correlation between age groups and HEV seroprevalence varies [13, 22]. The anti-HEV IgG seroprevalence among blood donors in the Qassim Region was comparable to that in Riyadh (8.4%) but was much lower than estimates for other regions of Saudi Arabia, such as Gizan (14.9%), Makkah (18.7%), and Jeddah (16.9%) [20-22]. Moreover, the seroprevalence of anti-HEV IgG was also lower than that reported in neighboring countries, such as Qatar (20.7%) [19]. This variation in the HEV seroprevalence among studies can be explained by differences in the commercial enzyme immunoassays used to detect HEV antibodies in blood donor samples. Commercial ELISA assays have different sensitivities and specificities, which may lead to variation in estimates of the HEV seroprevalence [23]. For instance, the HEV ELISA kit used in the present study (Fortress Diagnostics) has a sensitivity and specificity of 99.5% and 99.6%, respectively, but the most common ELISA for HEV detection (the Wantai assay) is known for its high antibody detection rates [24]. In addition, these assays may not detect all HEV genotypes. The variation among studies may also be explained by differences in demographic properties, which can vary among cities within the same country. For example, the higher HEV seroprevalence in Makkah and Jeddah compared to other cities in Saudi Arabia could be due to the high number of visitors of different nationalities in these two cities every year for pilgrimages and other religious practices. A potential limitation of the current study is that we could not confirm the presence of HEV RNA in the seropositive donor samples.

In summary, we showed that the seroprevalence of HEV is relatively low (5.7% and 1.3% for anti-HEV IgG and IgM, respectively) among blood donors in the Qassim Region, Saudi Arabia when compared to those in other regions of the country. The anti-HEV IgG seropositivity was significantly higher in non-Saudi donors (22.1%) than in Saudi donors (3%). Further investigations of the seroprevalence of HEV in a larger sample of blood donors from an expanded geographical distribution are needed to determine the need for HEV screening for blood donors in Saudi Arabia.

## Data Availability

All data are fully available without restriction.

## Acknowledgments

The authors gratefully thank all the staff in the Medical Laboratory Department at King Saud Hospital for their contribution to this study. We also would like to thank Editage (www.editage.com) for English language editing.

